# Fully automated histological classification of cell types and tissue regions of celiac disease is feasible and correlates with the Marsh score

**DOI:** 10.1101/2023.12.11.23299520

**Authors:** Michael Griffin, Aaron M. Gruver, Chintan Shah, Qasim Wani, Darren Fahy, Archit Khosla, Christian Kirkup, Daniel Borders, Jacqueline A. Brosnan-Cashman, Angie D. Fulford, Kelly M. Credille, Christina Jayson, Fedaa Najdawi, Klaus Gottlieb

## Abstract

**Aims:** Histological assessment is essential for the diagnosis and management of celiac disease. Current scoring systems, including modified Marsh (Marsh–Oberhuber) score, lack inter-pathologist agreement. To address this unmet need, we aimed to develop a fully automated, quantitative approach for histology characterisation of celiac disease.

**Methods:** Convolutional neural network models were trained using pathologist annotations of haematoxylin and eosin-stained biopsies of celiac disease mucosa and normal duodenum to identify cells, tissue and artifact regions. Human interpretable features were extracted and the strength of their correlation with Marsh scores were calculated using Spearman rank correlations.

**Results:** Our model accurately identified cells, tissue regions and artifacts, including distinguishing intraepithelial lymphocytes and differentiating villous epithelium from crypt epithelium. Proportional area measurements representing villous atrophy negatively correlated with Marsh scores (r=−0.79), while measurements indicative of crypt hyperplasia and intraepithelial lymphocytosis positively correlated (r=0.71 and r=0.44, respectively). Furthermore, features distinguishing celiac disease from normal colon were identified.

**Conclusions:** Our novel model provides an explainable and fully automated approach for histology characterisation of celiac disease that correlates with modified Marsh scores, facilitating diagnosis, prognosis, clinical trials and treatment response monitoring.

**KEY MESSAGES:** *What is already known on this topic:*

➢ Prior research has utilised machine learning (ML) techniques to detect celiac disease and evaluate disease severity based on Marsh scores.
➢ However, existing approaches lack the capability to provide fully explainable tissue segmentation and cell classifications across whole slide images in celiac disease histology.
➢ The need for a more comprehensive and interpretable ML-based method for celiac disease diagnosis and characterisation is evident from the limitations of currently available scoring systems as well as inter-pathologist variability.

*What this study adds:*

➢ This study is the first to introduce an explainable ML-based approach that provides comprehensive, objective celiac disease histology characterisation, overcoming inter-observer variability and offering a scalable tool for assessing disease severity and monitoring treatment response.

*How this study might affect research, practice or policy:*

➢ This study’s fully automated and ML-based histological analysis, including the correlation of Marsh scores, has the potential to enable more precise disease severity measurement, risk assessment and clinical trial endpoint evaluation, ultimately improving patient care.

## INTRODUCTION

Celiac disease, an autoimmune disease triggered by dietary gluten, affects around 1% of the population.^1^ Its diagnosis can be challenging due to symptom diversity, spanning from no symptoms to severe malabsorption.^1^ ^2^ Patients with celiac disease face a slightly increased overall risk of developing bowel lymphoma in comparison to the general population.^2^

Histological assessment is crucial for the diagnosis and management of celiac disease,^3^ as well as for endpoint assessment in clinical trials,^4^ with findings of intraepithelial lymphocytosis, crypt hyperplasia and villous atrophy indicative of the presence of the disease.^5^ Clinical study endpoints often rely on a quantification of disease activity, demonstrated by changes in histology and characterised according to disease severity by classification systems such as the modified Marsh (Marsh–Oberhuber) score.^3^ ^6^ However, inter-observer agreement is low for these metrics.^6^ The United States Food and Drug Administration recommends using a clinically accepted histological scale such as the Marsh score for screening samples in clinical studies of treatments for celiac disease, to ensure patient eligibility at enrolment. Furthermore, histology is also recommended as a co-primary endpoint in these studies.^7^

Celiac disease is often underdiagnosed due to variation between pathologists in their assessment of biopsy tissue samples,^8^ even if multiple biopsies are obtained.^3^ ^5^ Poor quality of biopsy tissue and overlapping histopathology features between related conditions may contribute to this variability.^5^ ^8^ ^9^ Recently, there has been increased interest in applying machine learning (ML) to pathology,^10^ ^11^ including to improve the accuracy and efficiency of celiac disease diagnosis.^12^

Such automation is expected to significantly reduce variability,^12^ ^13^ enabling smaller clinical studies to attain sufficient statistical power to demonstrate treatment effects. Indeed, convolutional neural network (CNN) tissue and cell model predictions from gastrointestinal samples have been used to create human interpretable features (HIFs) that enable the quantitative assessment of inflammatory pathological changes in non-celiac gastrointestinal diseases.^16^

While previous research has successfully employed ML to detect celiac disease and assess disease severity based on Marsh scores,^13^ this study aims to bridge critical gaps in the current research landscape. The work presented here represents the first report of an ML application for celiac disease that provides fully explainable tissue segmentation and cell classifications across whole slide images (WSIs) of duodenal mucosal biopsies. Through this approach, we have enabled the extraction of HIFs, such as cell densities, cell count proportions, and tissue area proportions, all of which exhibit correlations with Marsh scores. By utilising ML-based quantification, this study aims to objectively and exhaustively characterise celiac disease histology, address the limitations of manual histological assessments, and provide granular data for translational research and clinical trials. We believe such an approach has tremendous potential as a scalable tool for measuring disease severity and monitoring treatment response.

## MATERIALS AND METHODS

### Data set characteristics

WSIs of haematoxylin and eosin (H&E)-stained biopsies of duodenal mucosa of varying celiac disease severity (N=318) and mucosa of normal duodenum (N=58) were collected from PathAI Diagnostics (Memphis, USA) (**supplemental figure 1**).

The cohort size was determined based on the project’s scope and the availability of small intestine biopsies encompassing the full spectrum of celiac disease histology at the central laboratory. Slides were scanned at 40× objective magnification using the Aperio GT450 slide scanner (Leica Biosystems, Wetzlar, Germany). Celiac disease slides were split into training (n=230; 72.3%), validation (n=60; 18.9%) and test (n=28; 8.8%) datasets to ensure the even distribution of available patient metadata. For normal duodenum, slides were divided into a similar ratio of training (n=40; 69.0%), validation (n=12; 20.7%) and test (n=6; 10.3%) datasets.

### Machine learning-based tissue model development

We developed a model to identify and quantify relevant tissue regions, and we also utilised a previously trained model to identify and quantify cell types and artifact regions^16^ on H&E-stained WSIs of celiac disease and normal duodenum (**figure 1**). Using these identified cells and tissue regions, histological features relevant to celiac disease and representing surrogate measures of modified Marsh score components were quantified, including the proportion of intraepithelial lymphocytes to enterocytes in villous epithelium and the surface areas of villous epithelium and crypt epithelium. The latter two features assess villous height and crypt hyperplasia respectively.

**Figure 1.**
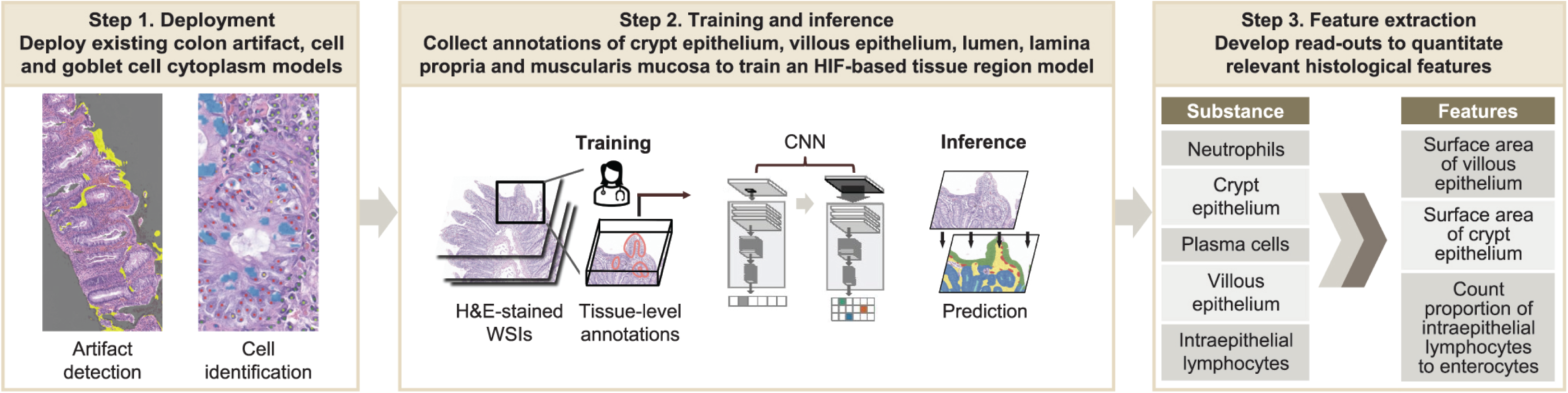
Proof-of-concept development of models based on HIFs on training data. CNN, convolutional neural network; H&E, haematoxylin and eosin; HIF, human interpretable feature; WSI, whole slide image.

WSIs were annotated by board-certified gastrointestinal pathologists. In total, 8356 tissue region annotations were collected. Annotations of crypt epithelium, villous epithelium, crypt lumen, lamina propria, blood vessels, muscularis mucosa and other tissue (including Brunner’s glands and submucosa) were used to train a HIF-based tissue segmentation region model. From these annotations, a CNN model was trained to produce pixel-level predictions of small intestinal mucosa tissue regions. Previously developed models to detect and exclude tissue artifacts and identify and classify the cells in colon tissue were also deployed.^16^ Tissue and cell model predictions were visualised as heatmaps on WSIs.

Heatmap transformations were used to remove artifact regions (e.g. debris, tissue folds, out-of-focus regions), extracting features only from high-quality tissue.

### Validation and review of cell and tissue models

A PathAI pathologist (F.N.) performed quality control of the tissue labels used for model training and qualitatively reviewed the tissue and cell overlays representing model predictions on H&E-stained WSIs. This qualitative review helped guide the iterative model development (**supplemental figure 2**).

To establish ground truth for cell model prediction accuracy, representative image frames were sampled (75 μm×75 μm; N=160). Frames were exhaustively annotated for all model-predicted cell types by five gastrointestinal pathologists. Hierarchical clustering was performed on these annotations and model predictions as previously described to identify cell locations.^16^ To account for potential pathologist bias and variability, Bayesian-estimated ground truths were used to quantify and compare the performance of the annotators and the model (**supplemental figure 3**).

### Evaluation of model-derived HIFs

HIFs (e.g. the proportional area of villous epithelium relative to lamina propria) were extracted from WSIs of normal duodenum (N=52) and scored celiac disease (N=118). HIFs were correlated with modified Marsh scores (type 0, normal lesions; type 1, infiltrative lesions; type 2, hyperplastic lesions; and types 3a, 3b and 3c, destructive lesions)^6^ using Spearman rank correlations. Scores only assessed the presence of >30 intraepithelial lymphocyte cells when differentiating scores 0 from 1 rather than quantifying any further increase in intraepithelial lymphocyte cells with increasing disease severity.

After establishing correlations between HIFs and modified Marsh scores, potential differences in the model-derived features between celiac disease and normal duodenum were evaluated.

### Statistical analysis

To assess cell model performance, the harmonised average of precision and sensitivity (F1 score) was calculated for both the cell model predictions and each pathologist annotation compared to the consensus on representative image frames. To evaluate the model-generated HIFs, each HIF was assessed for correlation with consensus modified Marsh scores using Spearman rank correlations. Data analyses in this study used the programming language Python (OpenEDG Python Institute, West Pomerania, Poland) for tissue and cell model development. Additionally, OpenSlide Python (Carnegie Mellon University, Pittsburgh, PA, USA) was used to load WSIs, Matplotlib (John D Hunter, Matplotlib Development Team and NumFOCUS, Austin, TX, USA) was used for plotting graphs, and PyTorch (PyTorch Foundation, the Linux Foundation, San Francisco, CA, USA) was used for tissue and cell model development.

To associate model-derived features of celiac disease following correlations with modified Marsh scores, mean (standard deviation) feature levels were used to show differences between celiac disease and normal duodenum. P values were calculated by independent *t*-test.

## RESULTS

### Model development for quantitation of celiac disease histological features

The tissue model developed, as well as the previously trained cell and artifact models,^16^ were deployed on H&E-stained WSIs of celiac disease and normal duodenum. Relevant cell types identified included neutrophils, plasma cells, enterocytes, intraepithelial lymphocytes, non-intraepithelial lymphocytes, eosinophils and goblet cells (**figure 2**); all other cell types are predicted as “other cells”. In addition, tissue regions identified included villous epithelium, crypt epithelium, lamina propria, muscularis mucosa and blood vessels (**figure 3**). Tissue regions such as total epithelium and mucosa could also be extracted from the tissue segmentation overlays. The tissue model distinguished villous epithelium from crypt epithelium.

**Figure 2.**
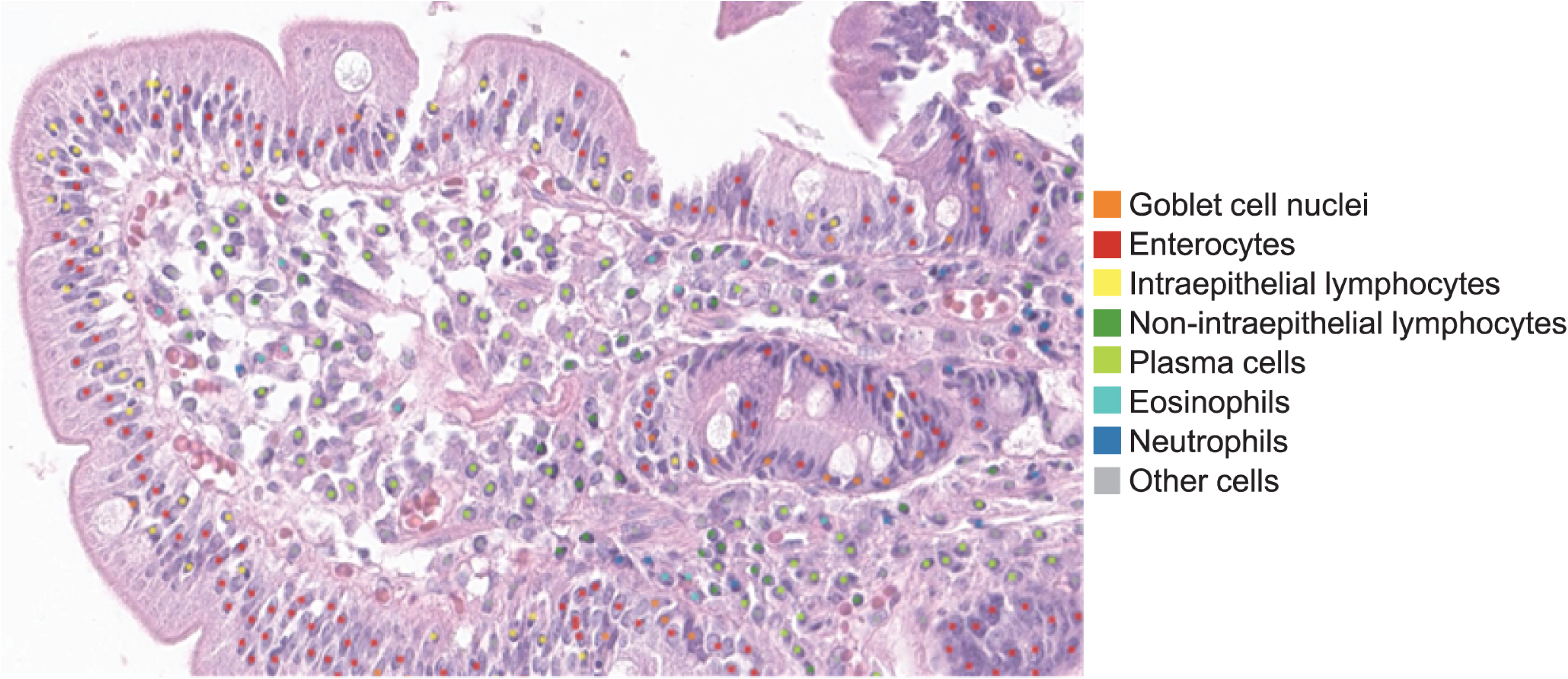
Overlays generated by cell segmentation model for model deployment.

**Figure 3.**
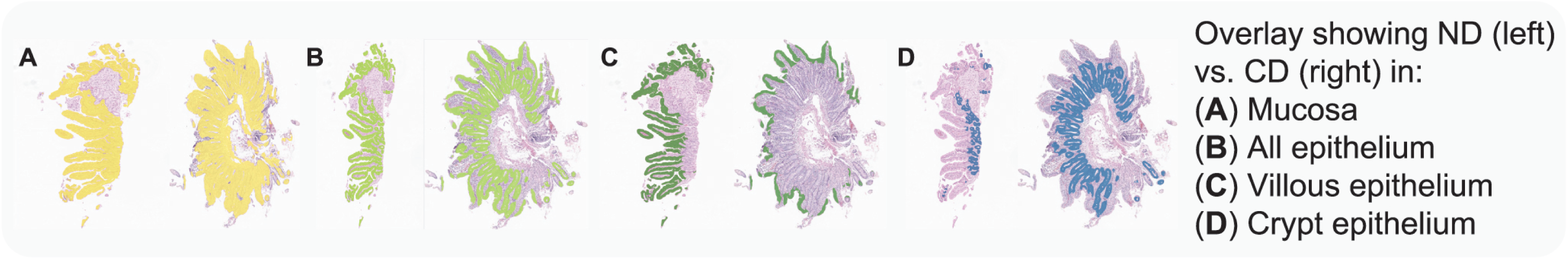
Tissue segmentation model showing distinct tissue regions. CD, celiac disease; ND, normal duodenum.

The cell model’s performance was validated by comparing it with pathologists’ annotations using Bayesian-estimated ground truths. Here, we sought to concentrate this validation on overlapping cells, focusing on cell confusion. The cell model demonstrated acceptable sensitivity for most cell types (**figure 4**).

**Figure 4.**
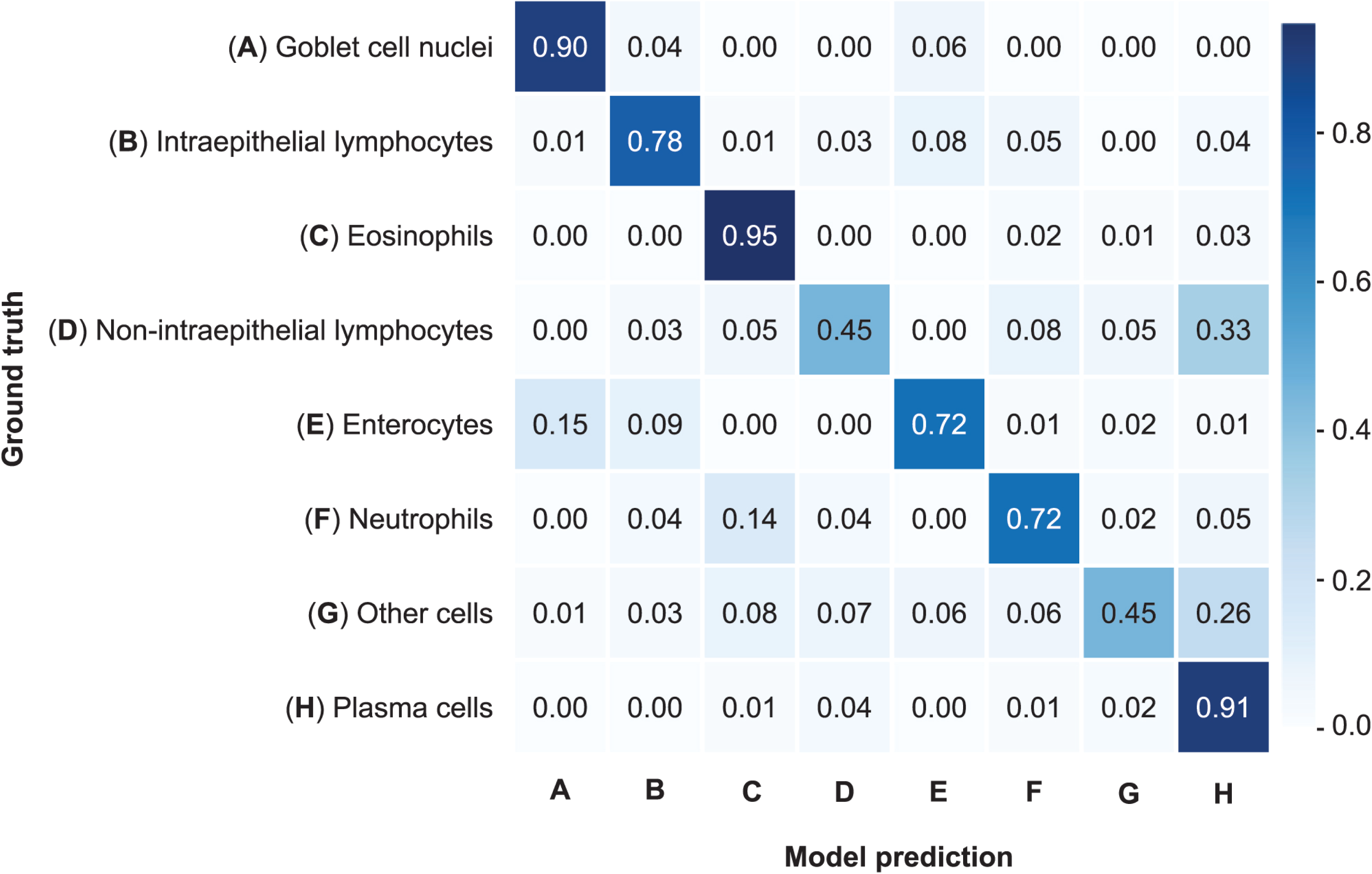
Cell model confusion matrix showing sensitivity across different cell types.

Cell model predictions were compared with labels from five gastrointestinal pathologists on representative image frames to determine model accuracy. We reported elements of the F1 score for both cell model predictions and pathologists’ labels for each of the cell types (**figure 5A**,**B**). Overall, cell model specificity remained relatively consistent and was similar to that of the pathologists for most cell types, with a slight difference being seen for plasma cells, while sensitivity was more variable outside the intraepithelial lymphocyte class.

**Figure 5.**
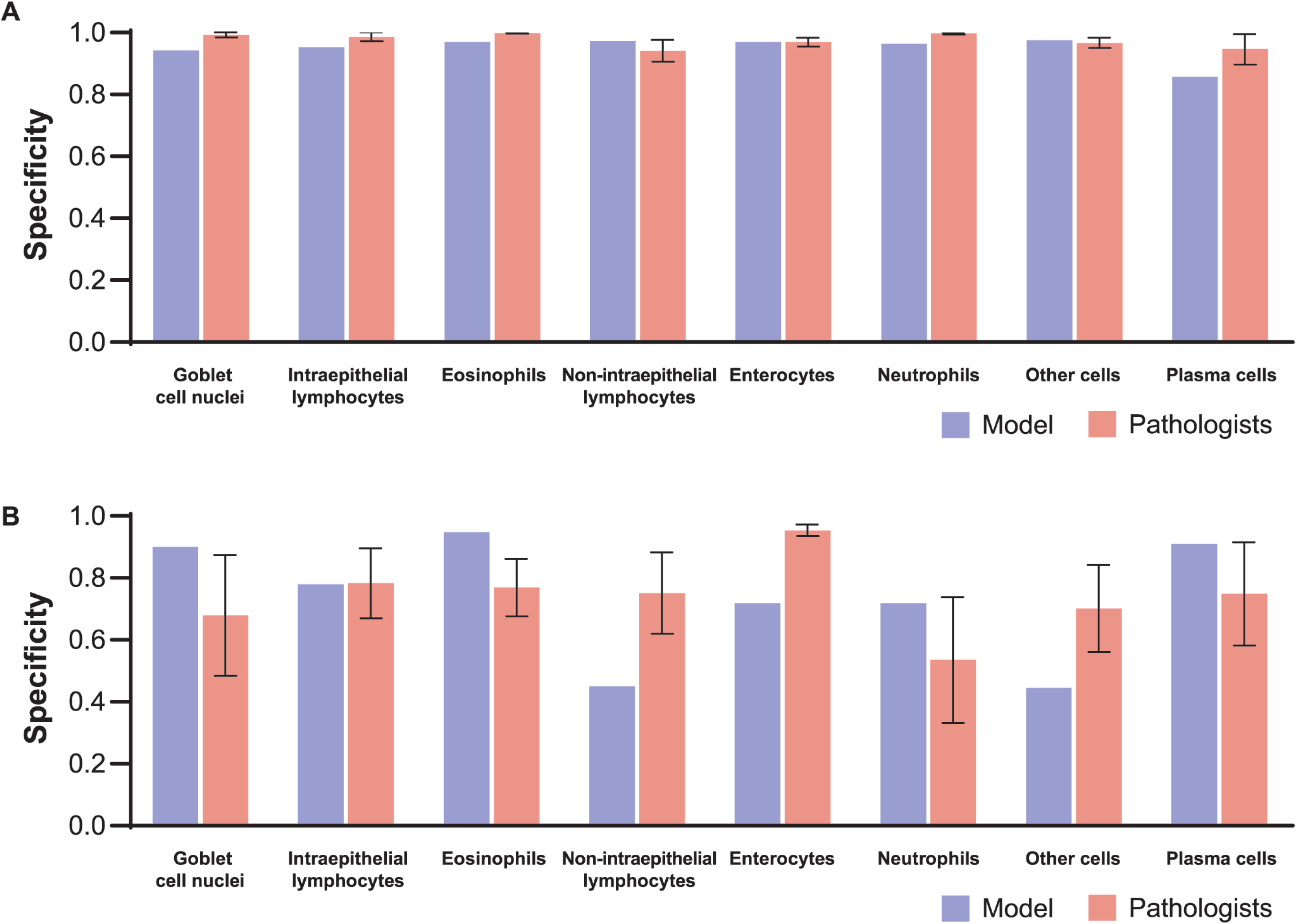
Accuracy of cell model predictions compared with pathologists. (A) Specificity comparison. (B) Sensitivity comparison.

### Correlation of surrogate features with modified Marsh score

HIFs from our models were analysed to assess correlation with modified Marsh scores. The area of villous epithelium relative to mucosa was negatively correlated with modified Marsh score (Spearman r=−0.79, p<0.0001) (**figure 6A**). The area of crypt epithelium in tissue (**figure 6B**) positively correlated with modified Marsh score (Spearman r=0.71, p<0.0001), as did the number of intraepithelial lymphocyte cells relative to enterocyte cells in villous epithelium (**figure 6C**) (Spearman r=0.44, p<0.0001). These results are summarised in **supplemental table 1**.

**Figure 6.**
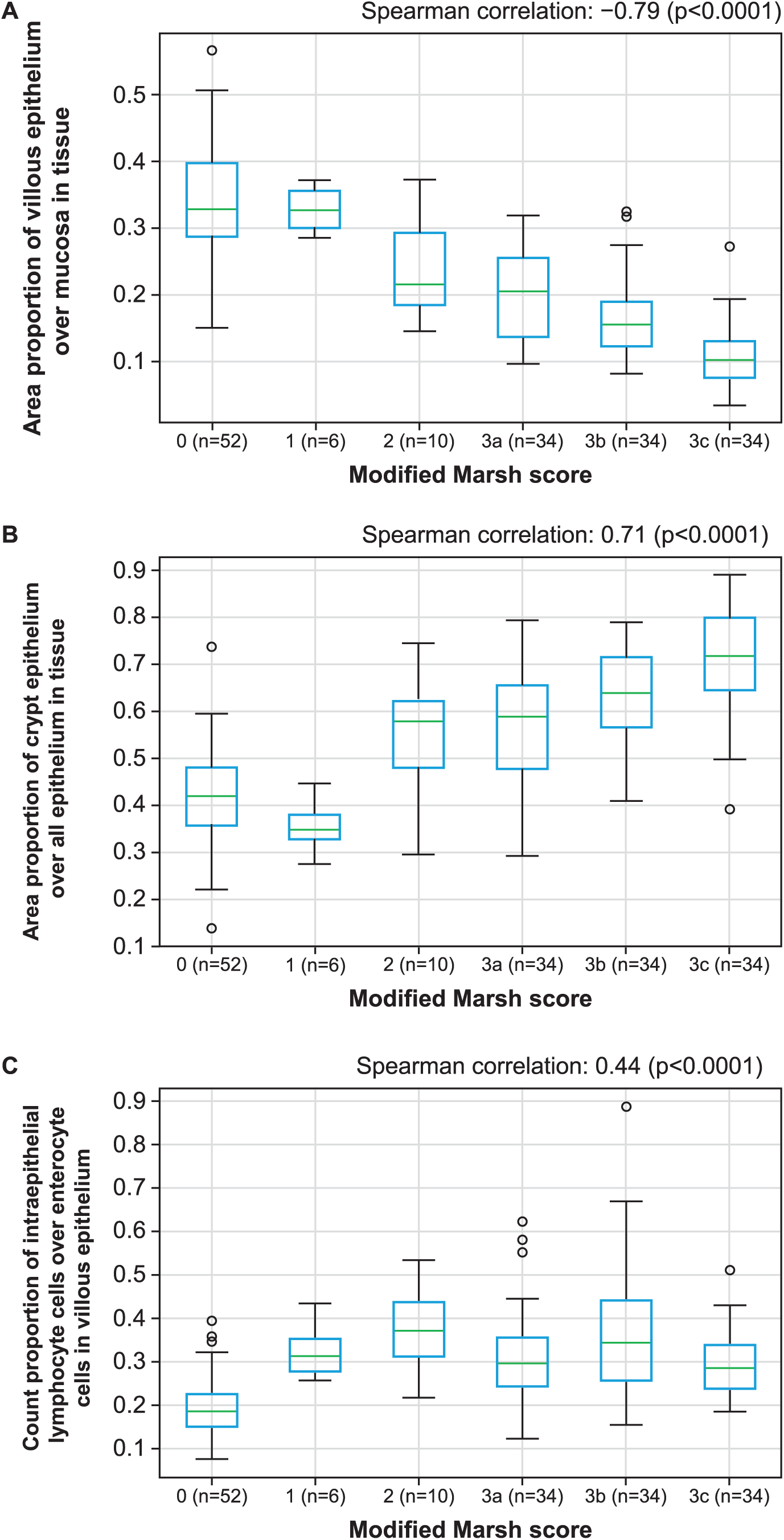
Example cell and tissue segmentation model correlation with modified Marsh score. (A) Surrogate features of villous blunting. (B) Surrogate features of crypt hyperplasia. (C) Surrogate features of intraepithelial lymphocyte infiltration.

The HIFs extracted from the cell and tissue models distinguished normal biopsies from those with celiac disease. For example, the proportional area of villous epithelium relative to mucosa and the proportional area of villous epithelium relative to crypt epithelium were both lower in celiac disease tissue compared with normal tissue, while the proportional area of crypt epithelium relative to total epithelium, the proportional area of lamina propria over mucosa and the density of intraepithelial lymphocytes in villous epithelium were higher in celiac disease (p<0.0001 for all comparisons) (**table 1**).

**Table 1.**
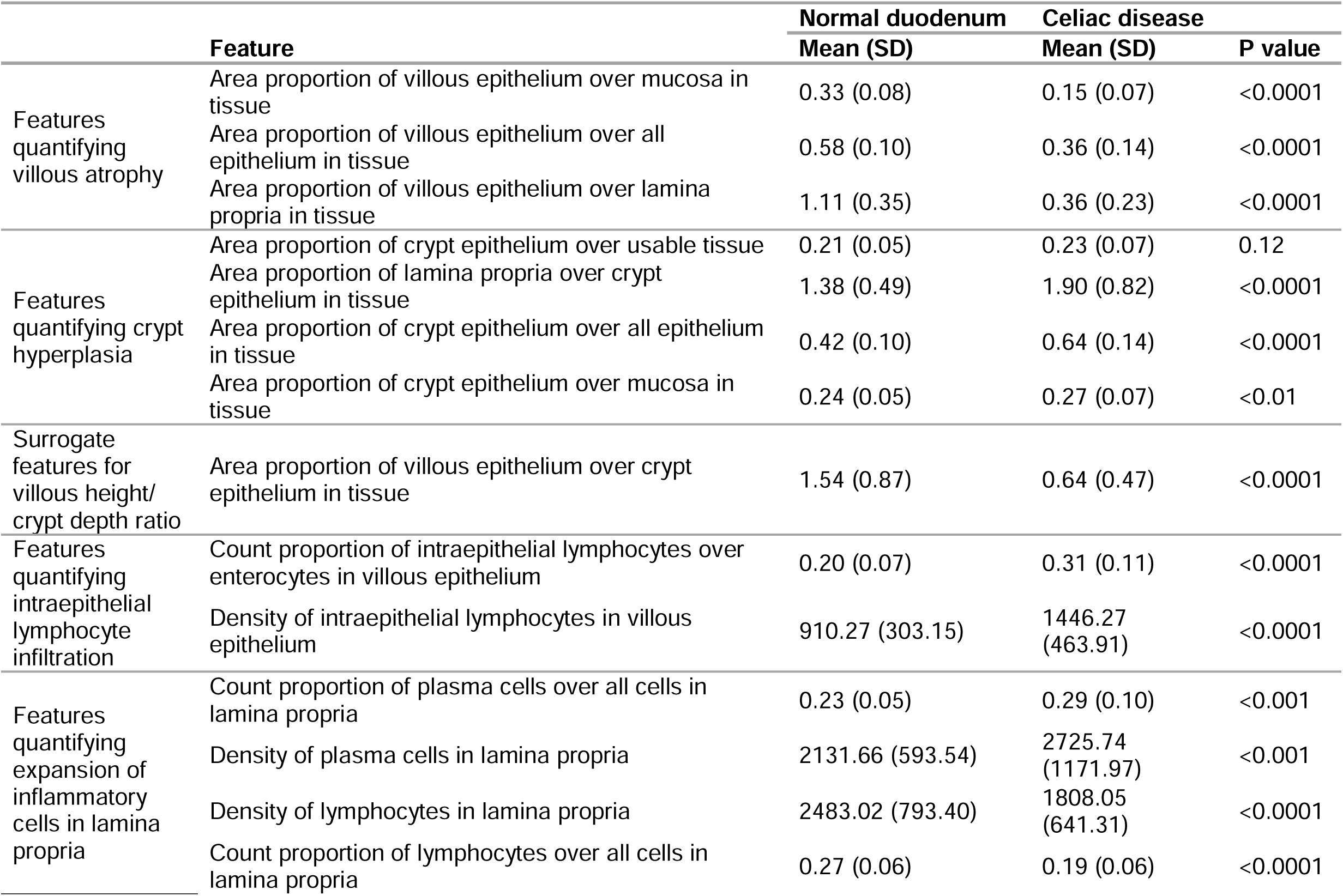
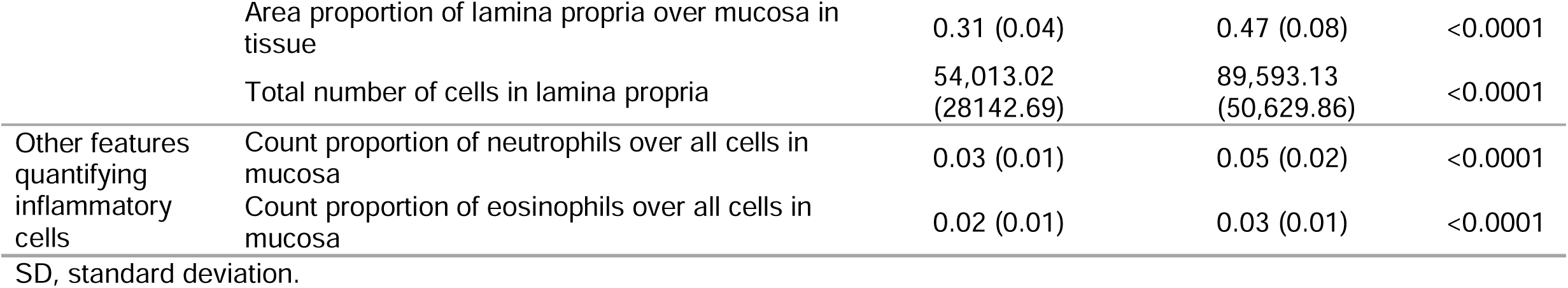
Association of model-derived features with celiac disease.

## DISCUSSION

Histological assessment of celiac disease plays a crucial role in diagnosing disease and evaluating the effectiveness of clinical interventions.^3^ However, inter-observer variability can affect the consistency and accuracy of results.^6^ To overcome this limitation and augment pathologists’ assessments of disease severity, we aimed to develop a fully automated and explainable approach to quantify the cellular and tissue-based features of celiac disease in H&E-stained clinical samples. The HIFs extracted from this model reflected histological changes that were measured by modified Marsh scores, potentially providing a quantitative and reproducible means to assess celiac disease severity.

Our model produced continuous feature measurements that can be interpreted as surrogate markers of celiac disease pathology (**supplemental table 1**). The relationship of these features with the ordinal Marsh score categories can be used as a benchmark to measure the model’s performance. For example, we examined the area of villous epithelium relative to the area of mucosa as an indicator of villous blunting, a hallmark of celiac disease, and found a negative correlation with higher modified Marsh scores. To gauge crypt hyperplasia, a more subtle feature, we examined the area of crypt epithelium relative to total epithelial area, revealing a positive correlation between this feature and Marsh scores at a Marsh score of 2 and above. The trained cell model directly quantitated the proportion of intraepithelial lymphocytes relative to the number of enterocytes within the villous structures. As expected, these values increased with disease severity.

Existing celiac disease scoring systems, such as the modified Marsh score, primarily rely on qualitative and descriptive categorisations, leading to subjectivity and limited sensitivity to subtle changes.^17^ In this study, we propose an alternative approach, utilising ML techniques to enable continuous, quantitative evaluation of the histological changes in celiac disease. By capturing histological alterations on a granular and objective scale, this novel approach offers enhanced sensitivity to changes in intraepithelial lymphocyte density, as well as villous and crypt epithelial surface area, overcoming limitations of the qualitative assessments of conventional manual scoring systems.

Some of our model-extracted HIFs directly quantified features of intraepithelial lymphocytes. This cell type is an essential consideration during disease assessment, as the presence of >30 intraepithelial lymphocytes per 100 enterocytes in the duodenum is a defining feature of celiac disease.^18^ The HIFs extracted from our model include count proportions and/or density of intraepithelial lymphocytes, specifically in the villous epithelium. This model also allowed for the extraction of features relating to intraepithelial lymphocytes in crypt epithelium and a comparison of their density in villous and crypt epithelium, providing a comprehensive overview of the spatial distribution of this cell type within distinct epithelial regions. Additional relevant features included the proportional area of villous epithelium (quantifying the change related to villous atrophy), the proportional area of crypt epithelium (quantifying crypt hyperplasia) and the ratio of villous epithelium area to crypt epithelium area (quantitatively capturing the relationship of villous height to crypt depth).^19^

The key strengths of this study become apparent when considering that these model-generated features not only bear relevance to the modified Marsh scoring system but are also essential components of the histological hallmarks of celiac disease (**table 1**).^19^ These HIFs encompass features not previously incorporated into any formalised scoring system, such as relative numbers and density of inflammatory cells (including lymphocytes, plasma cells, eosinophils and neutrophils) in lamina propria or in mucosa. These metrics characterise the immune micro-environment within celiac biopsies, as well as the total area and area proportion of lamina propria, capturing the expansion of lamina propria, a phenomenon known to be associated with disease activity.^19^

Furthermore, one of the key strengths of this study lies in our model’s capacity to discern between normal duodenum and celiac disease through the quantification of features associated with the disease microenvironment in mucosal biopsies. As expected, quantifying features of villous atrophy, as evidenced by reduced area proportion of this feature, and the augmented area proportion signifying mucosa crypt hyperplasia, distinguished between the histology of unaffected biopsies and those indicative of celiac disease. Supplementary quantitative attributes of the inflammatory microenvironment known to be associated with celiac disease, encompassing the infiltration of chronic inflammatory cells like lymphocytes and plasma cells within the lamina propria, coupled with the associated expansion of this layer,^19^ further distinguished normal biopsy samples from those with celiac disease.

Discernible differences between the two groups were also observed in the quantitative evaluation of granulocytes, which has been previously described.^20^ ^21^

While our model was limited by the small sample size, additional assessment involving larger cohorts will allow future refinement of the model’s performance. The cell model can also be trained specifically on duodenum biopsies and expanded to predict features associated with additional cell types (e.g. Paneth cells). An additional limitation of the current approach is related to the extraction of HIFs across a specific tissue area in the entire slide, which overlooks the potential variation between different tissue fragments. In a manual assessment of celiac disease in biopsies, pathologists often determine disease severity based on the most severely affected tissue region. To address this limitation, future work will focus on reporting HIFs separately for specific regions of interest within the tissue sample. This strategy is expected to allow for a more comprehensive and accurate assessment of disease severity within distinct tissue regions.

We foresee that ML-supported histological analysis will play a pivotal role in the advancement of precision medicine for patients with celiac disease. To our knowledge, this is the first report of fully explainable ML-based tissue and cell classifications across the WSIs of mucosal biopsies in celiac disease, enabling the extraction and statistical analysis of HIFs to empower translational research and clinical trials. The resulting quantitative model-generated HIFs can be used to build predictive models of existing Marsh scores or function as a continuous measurement, tracking histological change in celiac biopsies. Expanding upon this foundation, as we proceed to develop classification models aimed at predicting clinical outcomes alongside slide-level scores, we anticipate that the interpretability enabled by the utilisation of HIFs is poised to serve a dual purpose: validating the integrity of these models and revealing novel insights into disease biology. We believe that this ML-based assessment has tremendous potential as a scalable tool for measuring disease severity, risk stratification, prognostic evaluation, evaluating endpoints in clinical trials and monitoring of treatment responses; ultimately, advancing the care of patients with celiac disease.

## Data Availability

Model parameters for cell and tissue models, and codes for model training, inference and feature extractions are not disclosed. Access requests for such code will not be considered to safeguard PathAI's intellectual property. All feature tables, as well as source code, for reproducing correlational analyses will be deposited to GitHub prior to publication, and the link will be provided at that time. Access to cell- and tissue-type heatmaps, as well as usage of cell- and tissue-type classification models, are available on reasonable request to academic investigators, without relevant conflicts of interest, for non-commercial use who agree not to distribute the data. Access requests can be made to.

## Acknowledgements

The authors would like to thank all study participants. The authors are grateful to the software engineering and machine learning teams at PathAI for developing the systems and pipelines used for model development and feature extraction.

## Contributors

MG: conceptualisation, data curation, formal analysis, investigation, methodology, project administration, supervision, validation and visualisation, AMG: funding acquisition and writing (review and editing), CS: conceptualisation, data curation, formal analysis, investigation, methodology, validation and visualisation, QW: conceptualisation, data curation, formal analysis, investigation, methodology, validation and visualisation, DF: data curation, AK: conceptualisation, investigation, methodology, software and writing (review and editing), CK: conceptualisation, data curation and writing (review and editing), DB: conceptualisation, investigation, formal analysis and writing (review and editing), JABC: writing (original draft) and writing (review and editing), CJ: conceptualisation, data curation, formal analysis, investigation, methodology, project administration, validation and visualisation, FN: conceptualisation, data curation, formal analysis, investigation, methodology, project administration, validation, visualisation, writing (original draft) and writing (review and editing), KG: conceptualisation, funding acquisition, supervision and writing (review and editing). All authors: final approval of the manuscript. FN and KG are joint last authors.

## Patient consent for publication

Not required.

## Ethics approval

WCG IRB protocol number: 1316112

## REFERENCES

1 Lebwohl B, Sanders DS, Green PHR. Coeliac disease. Lancet 2018;391:70–81.

2 Marafini I, Monteleone G, Stolfi C. Association between celiac disease and cancer. Int J Mol Sci 2020;21:4155.

3 Rubio-Tapia A, Hill ID, Kelly CP, et al. ACG clinical guidelines: diagnosis and management of celiac disease. Am J Gastroenterol 2013;108:656–76.

4 Gottlieb K, Dawson J, Hussain F, et al. Development of drugs for celiac disease: review of endpoints for phase 2 and 3 trials. Gastroenterol Rep (Oxf) 2015;3:91–102.

5 Green PH, Cellier C. Celiac disease. N Engl J Med 2007;357:1731–43.

6 Corazza GR, Villanacci V, Zambelli C, et al. Comparison of the interobserver reproducibility with different histologic criteria used in celiac disease. Clin Gastroenterol Hepatol 2007;5:838–43.

7. US Food & Drug Administration (FDA). Celiac disease: developing drugs for adjunctive treatment to a gluten-free diet. 2022. https://www.fda.gov/regulatory-information/search-fda-guidance-documents/celiac-disease-developing-drugs-adjunctive-treatment-gluten-free-diet (accessed May 2023).

8 Mubarak A, Nikkels P, Houwen R, et al. Reproducibility of the histological diagnosis of celiac disease. Scand J Gastroenterol 2011;46:1065–73.

9 Syed S, Ehsan L, Shrivastava A, et al. Artificial intelligence-based analytics for diagnosis of small bowel enteropathies and black box feature detection. J Pediatr Gastroenterol Nutr 2021;72:833–41.

10 Rakha EA, Toss M, Shiino S, et al. Current and future applications of artificial intelligence in pathology: a clinical perspective. J Clin Pathol 2021;74:409–14.

11 Harrison JH, Gilbertson JR, Hanna MG, et al. Introduction to artificial intelligence and machine learning for pathology. Arch Pathol Lab Med 2021;145:1228–54.

12 Wei JW, Wei JW, Jackson CR, et al. Automated detection of celiac disease on duodenal biopsy slides: a deep learning approach. J Pathol Inform 2019;10:409–14.

13 Koh JEW, De Michele S, Sudarshan VK, et al. Automated interpretation of biopsy images for the detection of celiac disease using a machine learning approach. Comput Methods Programs Biomed 2021;203:106010.

14 Syed S, Al-Boni M, Khan MN, et al. Assessment of machine learning detection of environmental enteropathy and celiac disease in children. JAMA Netw Open 2019;2:e195822.

15 Sali R, Ehsan L, Kowsari K, et al. CeliacNet: celiac disease severity diagnosis on duodenal histopathological images using deep residual networks. Proceedings (IEEE Int Conf Bioinformatics Biomed) 2019;2019:962–67.

16 Najdawi F, Sucipto K, Mistry P, et al. Artificial intelligence enables quantitative assessment of ulcerative colitis histology. Mod Pathol 2023;36:100124.

17 Adelman DC, Murray J, Wu TT, et al. Measuring change in small intestinal histology in patients with celiac disease. Am J Gastroenterol 2018;113:339–47.

18 Bao F, Green PH, Bhagat G. An update on celiac disease histopathology and the road ahead. Arch Pathol Lab Med 2012;136:735–45.

19 Dickson BC, Streutker CJ, Chetty R. Coeliac disease: an update for pathologists. J Clin Pathol 2006;59:1008–16.

20 Moran CJ, Kolman OK, Russell GJ, et al. Neutrophilic infiltration in gluten-sensitive enteropathy is neither uncommon nor insignificant: assessment of duodenal biopsies from 267 pediatric and adult patients. Am J Surg Pathol 2012;36:1339–45.

21 Brown IS, Smith J, Rosty C. Gastrointestinal pathology in celiac disease: a case series of 150 consecutive newly diagnosed patients. Am J Clin Pathol 2012;138:42– 9.

