## Supplemental for "Fully automated histological classification of cell types and tissue regions of celiac disease is feasible and correlates with the Marsh score"

### TITLE

Fedaa Najdawi

PathAI, Inc.

1325 Boylston Street, Suite 10000

Boston, MA 02215

USA

### LIST OF TABLES AND FIGURES

**Supplemental Table 1** Correlations of model-derived HIFs with modified Marsh score

**Supplemental Figure 1** Workflow diagram of model development

**Supplemental Figure 2** Data quality control and qualitative review of tissue and cell models

**Supplemental Figure 3** Performance of cell predictions (frames) by the cell model

**Supplemental Table 1** **Correlations of model-derived HIFs with modified Marsh score**

| Feature | Spearman correlation coefficient | P value |
|---|---|---|
| Area proportion of villous epithelium over lamina propria in tissue | −0.836106 | <0.0001 |
| Area proportion of villous epithelium over mucosa in tissue | −0.788289 | <0.0001 |
| Area proportion of villous epithelium over all epithelium in tissue | −0.707178 | <0.0001 |
| Area proportion of villous epithelium over crypt epithelium in tissue | −0.707178 | <0.0001 |
| Area proportion of crypt epithelium over all epithelium in tissue | 0.707178 | <0.0001 |
| Area proportion of lamina propria over mucosa in tissue | 0.805047 | <0.0001 |
| Count proportion of intraepithelial lymphocytes over enterocytes in villous epithelium | 0.433375 | <0.0001 |
| Density of intraepithelial lymphocytes in villous epithelium | 0.527404 | <0.0001 |
| Density of plasma cells in mucosa | 0.587203 | <0.0001 |
| Count proportion of plasma cells over all cells in mucosa | 0.586244 | <0.0001 |
| Count proportion of eosinophils over all cells in mucosa | 0.594639 | <0.0001 |
| Density of eosinophils in mucosa | 0.612484 | <0.0001 |
| Count proportion of lymphocytes over plasma cells in mucosa | −0.639687 | <0.0001 |

HIF, human interpretable feature.

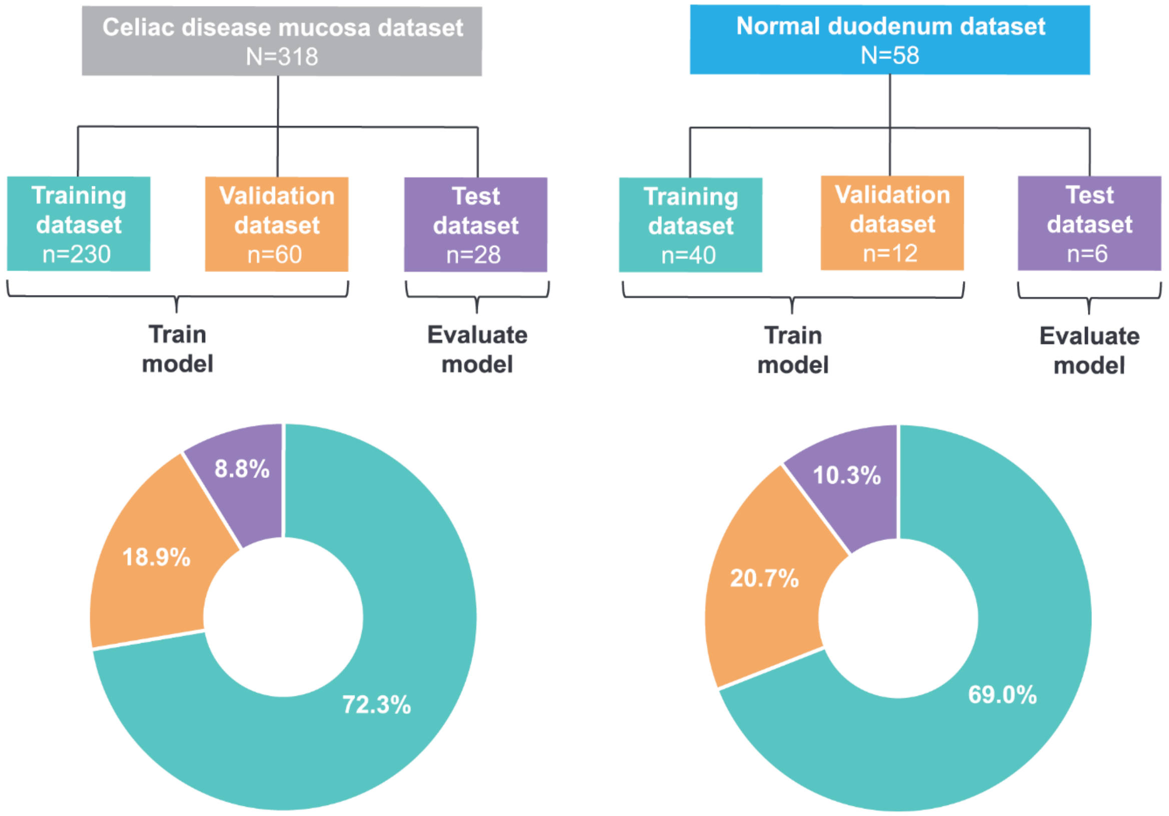


**Supplemental Figure 1** Workflow diagram of model development.

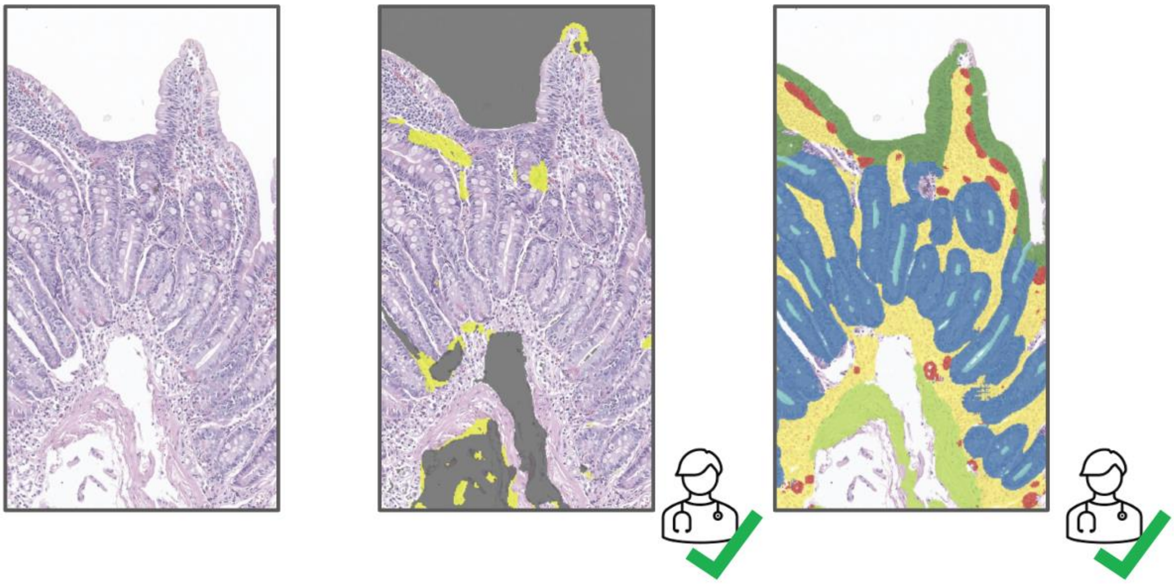

**Supplemental Figure 2** Data quality control and qualitative review of tissue and cell models.

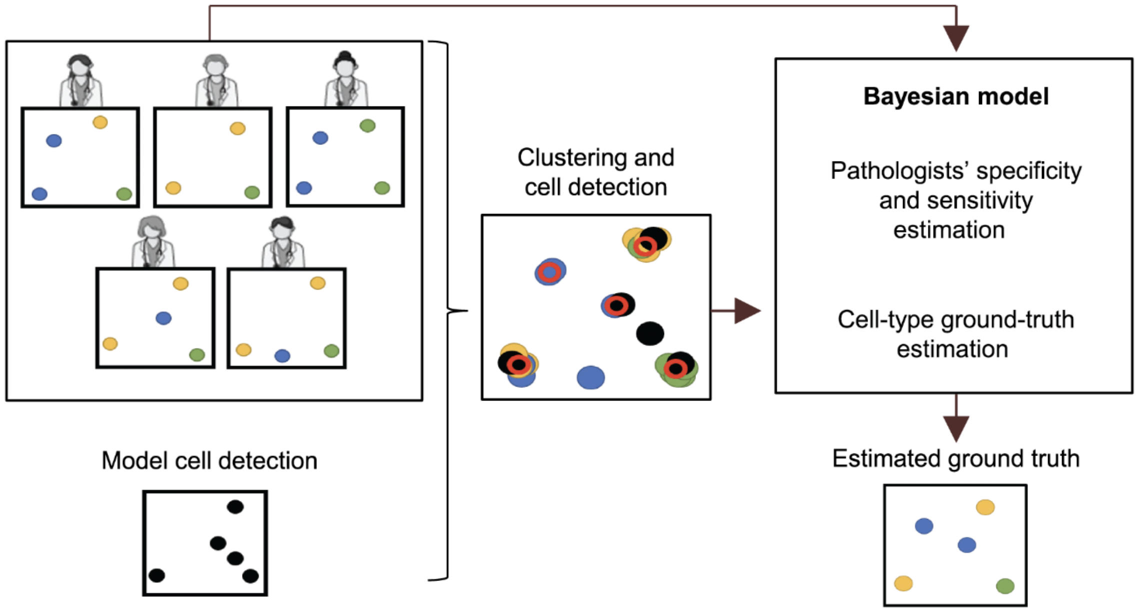


**Supplemental Figure 3** Performance of cell predictions (frames) by the cell model. Hierarchical clustering was performed on 1) annotations collected by pathologists (n=5) and 2) cell locations predicted by the cell model to locate true cells. A Bayesian model was then run using pathologist annotations as the input to estimate the ground truth cell type for each of the cell locations based on the estimated specificity and sensitivity of each annotator. Model predictions and pathologist predictions are then compared to that ground-truth.
